# Frontolimbic brain volume abnormalities in bipolar disorder with suicide attempts

**DOI:** 10.1101/2020.05.07.20093245

**Authors:** Mbemba Jabbi, Wade Weber, Jeffrey Welge, Fabiano Nery, Maxwell Tallman, Austin Gable, David E. Fleck, Elizabeth T.C. Lippard, Melissa DelBello, Caleb Adler, Stephen M. Strakowski

**Affiliations:** Department of Psychiatry, Dell Medical School, the University of Texas at Austin; The Mulva Clinic for the Neurosciences, Dell Medical School, the University of Texas at Austin; Institute of Neuroscience, the University of Texas at Austin; Department of Psychology, the University of Texas at Austin; Department of Psychiatry and Behavioral Neuroscience, University of Cincinnati College of Medicine, Cincinnati, OH

**Keywords:** Bipolar I Disorder, Mood Status, Suicide Attempt, Gray Matter Volume, Brain, Behavior

## Abstract

Over 2.3 million people in the United States live with bipolar disorder. Sixty percent of those with a bipolar disorder diagnosis attempt suicide at least once in their lifetime and up to 19% die by suicide. However, the neurobiology of suicide attempts in bipolar disorder remains unclear. We studied the gray matter volume (GMV) of 81 participants with a bipolar-I diagnosis (age-range: 14-34 years old) and 40 healthy participants (age-range 14.7-32 years old) to compare their neuroanatomy and histories of suicide attempt. In the bipolar group, 42 were manic with ages ranging from 14-30.6 years, and 39 were depressed with ages ranging from 14-34.3 years). Twenty three bipolar participants had a suicide attempt history, and 58 had no suicide attempt history. All participants completed behavioral/diagnostic assessments and MRI. We focused on a predefined frontolimbic circuitry in bipolar disorder versus controls to first identify diagnostic GMV correlates and to specifically identify GMV correlates for suicide attempt history. We found reduced GMV in bipolar diagnosis versus controls in the subgenual cingulate and dorsolateral prefrontal cortices. Our observed regional GMV reductions were associated with histories of suicide attempts and measures of individual variations in current suicidal ideation at the time of scanning.

**Highlights:** Suicide is a significant health problem, especially in bipolar disorder, but the neurobiological basis for suicide attempts remains obscure. We found that GMV reduction in anterior cingulate and dorsolateral prefrontal cortex associated with bipolar disorder, and suicide attempt history also correlated with suicidal ideation scores.

## INTRODUCTION

Suicide attempts are highly prevalent in mood disorders in general (Kessler et al. 2005; Murray et al. 2012; AFSP.org; NIMH.gov; CDC.gov) and bipolar disorder in particular (Dilsaver et al. 1994; Chen & Dilsaver 1996). Up to 60% of individuals with a diagnosis of bipolar disorder attempt suicide at least once in their lifetime (Dilsaver et al. 1994; Chen & Dilsaver 1996; Novick et al. 2010), and 10-15% of individuals with bipolar disorder die by suicide (Bostwick & Pankratz 2000). Despite that suicidal behaviors represent a significant public health problem, the neuroanatomical abnormalities underlying suicide attempt history (Bostwick & Pankratz 2000; Schmaal et al. 2020) and suicidal ideation in bipolar disorder are not well defined.

The frontolimbic network consisting of the anterior insula, dorsal anterior cingulate cortex (d-ACC), subgenual/ventral anterior cingulate cortex (vACC), the medial, orbitofrontal, and dorsolateral prefrontal cortices (DLPFC), are collectively identified to be critical for perceptual, mnemonic, experiential, and interoceptive aspects of affective processes (Dolan 2002; Damasio 2003; Critchley et al. 2004; Feinstein et al. 2006; Jabbi et al. 2008; Craig 2009). The ventral-to-dorsal extension of this frontolimbic brain circuitry, starting from the anterior insula sub-region extending to the ACC and prefrontal cortex via the uncinate fasciculus, is involved in regulating the integration of *<u>interoceptive</u>* (i.e., inner bodily feeling states) (Critchley et al. 2004; Craig et al. 2009; Joyce & Barbas et al. 2018) with *<u>exteroceptive</u>* (i.e., outer bodily percepts) (Nauta 1971; Goldman-Rakic 1988) sensory domains during affective processing. The integrity of this frontolimbic circuitry is suggested to contribute to the experience and maintenance of mood states (Khalsa et al. 2018) and suicidal behaviors (Wagner et al. 2012; Mathews et al. 2013; Oquendo et al. 2014; Lutz et al. 2017).

A preponderance of evidence points to structural magnetic resonance imaging (MRI) voxel-based morphometry (VBM) measures of reduced frontolimbic GMV in suicidal behavior in mood disorders (Eckman et al. 2010; Benedetti et al. 2011; Singh et al. 2012; Lijfijt et al. 2014; Huber et al. 2019; Fan et al. 2019; Lippard et al. 2019; Johnston et al. 2019; Schmaal et al. 2020). In bipolar I disorder in particular, frontolimbic GMV reductions in adults with a history of suicide attempts (Benedetti et al. 2011; Gosnell et al. 2016; Schmaal et al. 2020) are associated with the lethality of the attempts (Duarte et al. 2017). However, the relationship between these frontolimbic GMV neuroanatomical abnormalities with suicide attempt history and current suicidal ideations in bipolar disorder remains obscure. Lack of a better understanding of the neuroanatomical link between suicide attempt history and suicidal ideations impedes our understanding of brain anatomical abnormalities underlying suicidal risk behaviors. In this study, we specifically examined the neuroanatomical basis in terms of GMV correlates for suicide attempt history and active suicidal ideation in bipolar disorder.

Based on prior imaging evidence, we tested the hypothesis that frontolimbic GMV diagnostic markers for bipolar I disorder vs. healthy participants would show greater abnormalities with a bipolar suicide attempt history relative to no history of a suicide attempt. We further evaluated whether neurobiological correlates for a history of a suicide attempt would be associated with mood state and assessed if the suicide history-related GMV measures were associated with more acute suicidal ideation scores.

## METHODS

### Participants

All participants in this analysis completed behavioral, diagnostic, and structural MRI procedures following the design of the completed Bipolar Imaging and Treatment Research Center (BITREC) study of the University of Cincinnati; details of inclusion and exclusion criteria are available in previous reports (Borgelt et al. 2018; Fleck et al. 2019; Nery et al. 2019). One hundred and twenty-one participants were included in the final analysis of this structural MRI study, of which 42 were unmedicated bipolar I manic (11 with suicide attempt history), 39 unmedicated bipolar I depressed participants (12 with suicide attempt history), and 40 healthy comparison participants (see **Table 1** for demographic details and specific mood state as well as suicidal ideation ratings). For this analysis, we combined the mood states based on the assumption that neuroanatomy would minimally change in different phases of illness. All adult participants consented (all minor participants completed parental consent and child assents) to voluntarily participate in the study with the option to withdraw at any time of their choosing. All participants received compensation for participating in the study, and the protocol was approved by the University of Cincinnati IRB. In total, 12 bipolar manic, 5 bipolar depressed, and 1 control participant’s MRIs scans were excluded from the analysis (these excluded individuals were not part of the final 121 participant data that was analyzed) because of excessive movement. Furthermore, 4 bipolar manic and 2 bipolar depressed were not included in the final analysis of the reported 121 participants because of missing suicide attempt history information.

**Table 1.** Demographics and Diagnostics of Participant Samples. Table 1 shows data for all included participants in terms of mean age and mean YMRS, HAMD, and HMAD-suicidal ideation self-ratings and standard deviations (SD) per group (i.e., healthy controls, and the bipolar illustrated in the two subgroups of manic and depressed). Abbreviations: SD, standard deviation; No SA, no suicide attempt history; SA, suicide attempt history.

| TABLE 1 |  |  |  |  |  |  |
| --- | --- | --- | --- | --- | --- | --- |
| DIAGNOSTIC & BIPOLAR MOOD STATE DEMOGRAPHICS |  |  |  |  |  |  |
|  | HEALTHY - All Useable Scans (one per participant) |  |  |  |  |  |
|  | N (% of group) | Mean Age (SD) | (Females) | Mean YMRS (SD) | Mean HAMD (SD) | Mean HAMD Suicide Item (SD) |
| **Total** | 40 (33.05% study sample) | 22.45 (5.44) | 21 | 0.58 (1.32) | 1 (1.88) | 0 (0) |
|  | MANIC - All Useable Scans (one per participant) |  |  |  |  |  |
|  | N | Mean Age (SD) | (Females) | Mean YMRS (SD) | Mean HAMD (SD) | Mean HAMD Suicide Item (SD) |
| **Total** | 42 (34.7% study sample) | 18.45 (4.68) | 27 | 24.1 (4.89) | 15.48 (8.43) | 0.4 (0.88) |
| **No SA Attempt** | 31 (73.8% manic sample) | 19.28 (5.13) | 19 | 24.39 (4.86) | 15.16 (8.25) | 0.35 (0.19) |
| **SA Attempt** | 11 (26.2% manic sample) | 16.1 (1.53) | 8 | 23.27 (5.12) | 16.36 (9.28) | 0.55 (0.82) |
|  | DEPRESSED - All Useable Scans (one per participant) |  |  |  |  |  |
|  | N | Mean Age (SD) | (Females) | Mean YMRS (SD) | Mean HAMD (SD) | Mean HAMD Suicide Item (SD) |
| **Total** | 39 (32.23% study sample) | 23.43 (5.59) | 26 | 13 (6.50) | 27.54 (8.47) | 0.62 (.935) |
| **No SA Attempt** | 27 (69.2% depressed sample) | 23.13 (5.02) | 17 | 12 (5.94) | 25.81 (8.67) | 0.52 (0.89) |
| **SA Attempt** | 12 (30.8% depressed sample) | 24.1 (6.89) | 9 | 13 (7.87) | 31.42 (6.81) | 0.83 (1.02) |

### Behavioral Analysis

Demographics (sex and age), behavioral, diagnostic measures assessing IQ, bipolar I mood episode status of depression vs. mania, and individual ratings of mania, as measured with the Young Mania Rating Scales (YMRS) (Young et al. 2000), and depression, as measured with the Hamilton depression rating scale (HAM-D) (Hamilton 1960), were obtained at the time of scanning. Age of bipolar onset, mixed episode status, length of mood episode at the time of MRI scanning in weeks, and education in years was assessed. Drug abuse history was assessed as an exclusion criterion. Alcohol use (i.e., #of pints drank in past 30 days), cannabis use (i.e., #of times a cannabis cigarette was smoked in past 30 days), cigarette smoking (i.e., #of packs/day in past 30 days) were assessed, and group differences analyzed together with the age of disease onset, mixed episodic status, mood ratings, and demographics. Suicide attempt history was also assessed before MRI.

### Suicide Attempt Assessments

Data on participant suicide attempt status were retrieved from diagnostic interview records. Specifically, scores were extracted from the Washington University Kiddie Schedule for Affective Disorders and Schizophrenia (WASH-U KSADS) for participants younger than the age of 18 years (Kaufman et al. 2004); and the Structured Clinical Interview for DSM-IV (SCID) (DSM IV 1994) for adults. From these interviews, the presence of a suicide attempt history was extracted from interviewer notes. All healthy participants were free of any mental illness or suicidal behaviors. Additionally, the current and past Major Depressive Episode sections, which provide prompts to probe for suicidality, contained additional information for participants who responded with previous/current suicidal ideation or attempts. To specifically assess suicidal ideation, we used the HAM-D suicidal ideation item scores.

### Image Acquisition and Preprocessing

#### Anatomical Image Acquisition

All scans were performed on a 4 Tesla Varian Unity INOVA Whole Body MRI/MRS system (Varian Inc., Palo Alto, CA, USA) at the University of Cincinnati’s Center for Imaging Research. High-resolution, T1-weighted, 3-D neuroanatomic scans were acquired using a modified driven equilibrium Fourier transform (MDEFT) pulse sequence (TMD = 1.1 s, TR/TE = 13/6 ms, flip angle = 22°, FOV = 256 x 192 x 192 mm, matrix 256 x 192 x 96 pixels) as previously described (Lee et al., 2012).

#### Anatomical Image Preprocessing

The anatomical scans were processed using the Computational Anatomy Toolbox (CAT12) and the Statistical Parametric Mapping (SPM12) software. First, the scans were normalized using the Montreal Neurological Institute’s MNI152 template and then segmented according to tissue type (i.e., gray matter, white matter, cerebrospinal fluid, etc.). During the segmentation process, CAT12 also corrects for signal bias, noise, and global intensities. The segmented anatomical scan files were inspected for quality assurance, all scans below the 70% of acceptable quality assurance threshold defined prior to the preprocessing stage were excluded from the final analyzed sample (see participant section above for more details on the number and reasons for exclusion). Additionally, any scan with excessive movement (indicated by a blurry scan) or artefacts present were removed to ensure data quality. Intracranial volume was extracted from the data and the final preprocessed data were then smoothed with a Gaussian kernel of 8 mm full width at half maximum.

### Analyses of Extracted Frontolimbic Region of Interest (ROI) GMV Differences

The frontolimbic regions of interest (ROI) consist of the anterior insula, dACC, vACC, and DLPFC, which were defined using ITK Snap software on a standardized (in MNI space) anatomical brain template (http://www.itksnap.org/pmwiki/pmwiki.php). For each region, the anatomical boundary was demarcated in 3-dimensional space (see **Figure 1A-D**).

**Figure 1.**
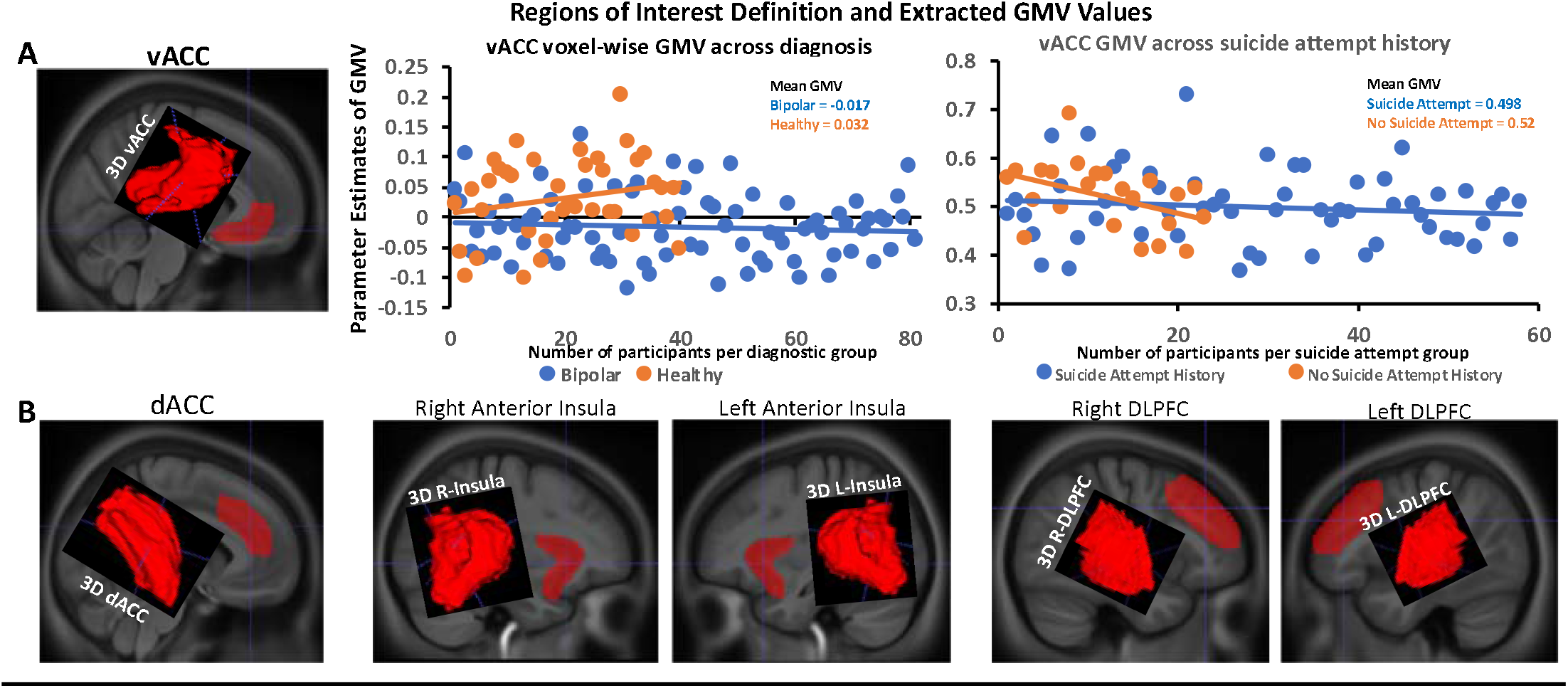
Regions of Interest Defined Volumes and ANOVAs. **A)** vACC region of interest anatomical demarcation (in red shown on the brain volume and in 3-dimensional view), and extracted individual GMV values from the same region showing scatter plots between healthy (lighter dots) > bipolar disorder (blue dots) as shown in the left top graph; as well as extracted individual GMV values from the same region showing a relatively more reduced GMV on average in those with no suicide attempt history (blue dots) vs. those with suicide attempt history (lighter dots) as shown in the right top graph. **B**) Remaining anatomical frontolimbic regions of interest that did not show significant GMV differences for diagnosis and suicide attempt history.

We then used Marsbar release version .44 to import individually defined frontolimbic ROIs from a cluster image (nifti), and then used the Marsbar function to extract GMV values from each ROI separately for each participant from the raw GMV values in native space (http://marsbar.sourceforge.net/). Because the participants include those below and above 18 years of age, age, sex, and total intracranial volume (TIV) were both included in a T-test as covariates when group differences were assessed in GMV (bipolar I vs. healthy) for each frontolimbic ROI to identify GMV group markers. Age, sex, and TIV were also included in a separate full factorial ANOVA as covariates in the model assessing suicide history-related differences in GMV markers (i.e., bipolar participants with suicide attempt history vs. bipolar participants with no suicide attempt history).

### Analyses of Frontolimbic-brain circuitry VBM of GMV Differences

Because the frontolimbic ROI approach was based strictly on anatomically defined regional boundaries that do not take into account the localized nature of GMV differences in specific sub-regions of the targeted frontolimbic circuitry, we additionally performed *post hoc* frontolimbic-wide GMV measures using voxel-based morphometry (VBM) in the Statistical Parametric Mapping (SPM12) software (https://www.fil.ion.ucl.ac.uk/spm/software/spm12/). As in the anatomical frontolimbic ROI analysis, age, sex, and TIV were included in a voxelwise T-test as covariates when group differences in GMV (bipolar I vs. healthy) were assessed in SPM12 to identify diagnostic markers. To this aim, we first evaluated the GMV differences between healthy participants (n=40) and the entire bipolar disorder group (n=81) to identify regional gray matter reductions associated with a bipolar I disorder diagnosis using a 2-sample T-test. We defined second-level contrasts derived from a random-effects analysis comparing healthy > bipolar GMV measures and bipolar > healthy groups at p < 0.001 uncorrected.

For our primary analysis, we included age, sex, and TIV in a frontolimbic circuitry voxelwise full-factorial ANOVA as covariates to assess GMV correlates for suicide attempt history (i.e., bipolar participants with suicide attempt history vs. bipolar participants without suicide attempt history). We adopted a similar approach for our *post hoc* analysis of GMV correlates for suicidal ideation scores by including individual-level scores of current suicidal ideation in a factorial model including depressed and manic bipolar participants with suicide attempt history and without suicide attempt history in SPM12. Contrasts of main effects of suicide attempt history across bipolar participants, as well as the main effect of suicide attempt history in relationship with current suicidal ideation scores, were examined separately in SPM12 at p < 0.001 uncorrected, using a frontolimbic mask (Supplementary **Figure 1**) for visualization.

## RESULTS

### General Linear Model (GLM) of Behavioral and Diagnostic/Mood Ratings

Sex and age distributions did not differ between bipolar I and healthy groups (F_1,119_ = 1.88; p=0.17). As expected, all mood ratings (HAM-D, YMRS, and the HAM-D item-11 suicidal ideation rating) were higher in the bipolar I group compared to the healthy comparison group (F_1,117_ < 10, p<0.001). Within the bipolar I groups (i.e., depressed versus manic), the manic group (mean age=18.45 ±.72 SEM) were significantly younger than the depressed group (mean age=23.43 ±.89 SEM) at F_1,77_ = 22.23 10, p<0.0001. By definition, overall HAM-D ratings of depressed mood were higher in the depressed than the manic group (F_1,80_ = 46.76; p=0.0001), while YMRS ratings of manic mood were higher in the manic than the depressed group (F_1,80_ = 25.50; p = 0.0001). At the time of scanning, the number of individuals experiencing a mixed-mood episode in the depressed group was also found to be higher than in the manic group (mean number of mixed episodes difference between the depressed and manic group = .36 ±.09 SEM). Furthermore, the age of bipolar disorder onset/age at first diagnosis was younger for the group with no suicide attempts (mean age of bipolar onset=15.01 ±.71 SEM) than the group with a suicide attempt history (mean age of bipolar onset=17.30 ±1.09 SEM) at F_1,76_ = 4.24, p<0.043.

We excluded all participants with a drug use/substance abuse history except for cannabis which was found to be more prevalent in the recruited population. Alcohol use, cigarette, and cigar smoking were not significantly different between the bipolar and healthy groups, between the bipolar mood states, or the suicide attempt status groups. Cannabis use was higher in the bipolar group than healthy comparisons but not between the bipolar mood states or suicide attempt status subgroups.

### GLM of Extracted Frontolimbic Region of Interest (ROI) GMV Measures

Frontolimbic ROI extractions of GMV values were found to be reduced in bipolar I vs. the healthy comparison group in the vACC (F_1,116_ = 7.37; p = 0.008) (**Figure 1A**), but not in any other ROI (**Table 2A**).

**Table 2.** ROI statistics of the observed GMV differences in Healthy > bipolar; and the main effects of suicide attempt history. Table 2 shows the region measures of MRI detected GMV differences in F-statics, Cohen’s d, and uncorrected p-values (Puncorr) for each brain region of interest in the healthy versus bipolar 2-sample t-test contrast (Table 2A), and in the main effect of suicide attempt factorial analysis (Table 2B). Abbreviations: ROI, the region of interest; vACC, ventral anterior cingulate cortex; dACC, dorsal anterior cingulate cortex; DLPFC, the dorsolateral prefrontal cortex.

| **Table 2A, ROI ANOVA, Healthy > Bipolar Disorder** |  |  |  |
| --- | --- | --- | --- |
| **ROI** | **F** | **Cohen's d** | **p<sub>uncorr</sub>** |
| vACC | 7.37 | 0.49 | 0.008 |
| dACC | 1.96 | 0.25 | 0.16 |
| left-Anterior Insula | 0.66 | 0.14 | 0.42 |
| right-Anterior Insula | 0.62 | 0.14 | 0.43 |
| left-DLPFC | 0.46 | 0.12 | 0.50 |
| right-DLPFC | 1.84 | 0.24 | 0.18 |
| **Table 2B, ROI ANOVA, Main Effect of Suicide Attempt** |  |  |  |
| **ROI** | **F** | **Cohen's d** | **p<sub>uncorr</sub>** |
| vACC | 74.09 | 1.91 | 0.047 |
| dACC | 3.5 | 0.41 | 0.06 |
| left-Anterior Insula | 0.07 | 0.05 | 0.78 |
| right-Anterior Insula | 2.43 | 0.34 | 0.12 |
| left-DLPFC | 0.02 | 0.03 | 0.9 |
| right-DLPFC | 0.31 | 0.12 | 0.58 |

Within the GMV-reduced vACC ROI found in our bipolar vs. healthy control comparison, our extractions of GMV values revealed a *main effect of suicide attempt* history in the bipolar disorder cohort (F_1,75_ = 4.09; p = 0.047) (**Figure 1A**). Specifically, the bipolar participants without suicide attempt history showed a more pronounced reduction in GMV relative to those with a history of a suicide attempt. This finding did not confirm our hypothesis. No other targeted ROIs (i.e., anterior insula, dACC, and DLPFC) showed diagnostic or suicide attempt history-related GMV differences (**Table 2B**). Extracted ROI values were not significantly correlated with HAM-D item-11 suicidal ideation scores.

### Voxelwise Frontolimbic Circuitry GMV Differences Between Bipolar vs. Healthy VBM

To first assess if measures of reduction in GMV within the anatomically defined frontolimbic ROIs might be more sub-regionally localized in relation to diagnostic group differences between bipolar I and healthy participants, we performed a whole-brain VBM 2-sample T-test comparison of GMV values. We found localized reductions of vACC/orbitofrontal cortex and bilateral DLPFC volumes (at p < 0.001 uncorrected, **Figure 2; Table 3A**) in the bipolar I group vs. healthy participants.

**Table 3.** Voxel-wise statistics of the observed GMV differences in Healthy > bipolar, and the full-factorial effect of suicide attempt history. Table 3a shows the whole-brain results of MRI detected GMV differences in Z-statics, Cohen’s d, and uncorrected p-values (Puncorr) for each brain region of interest in the healthy versus bipolar 2-sample t-test contrast (Table 3A), and the main effect of suicide attempt history (Table 3B); whereas Table3C shows the voxel-wise correlation between the right DLPFC GMV and measures of suicidal ideation in individuals with a suicide attempt history. Abbreviations: vACC, ventral anterior cingulate cortex; DLPFC, dorsolateral prefrontal cortex; X, Y, Z, and anatomical brain coordinates in Montreal Neurological Institute space; FEW, family-wise error corrected; FDR, false discovery rate corrected.

| **TABLE 3A Frontolimbic Voxel-wise Healthy > all Bipolar Disorder** |  |  |  |  |  |  |  |  |  |  |  |  |
| --- | --- | --- | --- | --- | --- | --- | --- | --- | --- | --- | --- | --- |
| Region | Position |  |  | Cluster-Level |  |  |  | Peak-Level |  |  |  |  |
|  | X | Y | Z | FWE | FDR | k<sub>E</sub> | p<sub>uncorr</sub> | FWE | FDR | Z | Cohen's d | p<sub>uncorr</sub> |
| vACC 1 | 3 | 14 | -8 | 0.15 | 0.53 | 472 | 0.02 | 0.25 | 0.32 | 4.13 | 0.78 | 0.000 |
| vACC 2 | 9 | 35 | -17 | 0.82 | 0.84 | 108 | 0.24 | 0.74 | 0.54 | 3.68 | 0.69 | 0.000 |
| vACC 3 | 8 | 44 | -8 | 0.99 | 0.90 | 5 | 0.83 | 0.99 | 0.92 | 3.12 | 0.58 | 0.001 |
| Right-DLPFC | 51 | 17 | 32 | 0.91 | 0.84 | 70 | 0.34 | 0.81 | 0.60 | 3.61 | 0.68 | 0.000 |
| Left-DLPFC | -35 | 8 | 36 | 0.61 | 0.80 | 183 | 0.13 | 0.46 | 0.32 | 3.92 | 0.74 | 0.000 |

| **TABLE 3B Frontolimbic Voxel-wise Full Factorial Main Effect of Suicide Attempt** |  |  |  |  |  |  |  |  |  |  |  |  |
| --- | --- | --- | --- | --- | --- | --- | --- | --- | --- | --- | --- | --- |
| Region | Position |  |  | Cluster-Level |  |  |  | Peak-Level |  |  |  |  |
|  | X | Y | Z | FWE | FDR | k<sub>E</sub> | p<sub>uncorr</sub> | FWE | FDR | Z | Cohen's d | p<sub>uncorr</sub> |
| Right-DLPFC | 57 | 23 | 27 | 0.99 | 0.88 | 18 | 0.57 | 0.98 | 0.78 | 3.39 | 0.83 | 0.000 |

| **TABLE 3C Frontolimbic Suicide Attempt-GMV Predicts Suicidal Ideation** |  |  |  |  |  |  |  |  |  |  |  |  |
| --- | --- | --- | --- | --- | --- | --- | --- | --- | --- | --- | --- | --- |
| Region | Position |  |  | Cluster-Level |  |  |  | Peak-Level |  |  |  |  |
|  | X | Y | Z | FWE | FDR | k<sub>E</sub> | p<sub>uncorr</sub> | FWE | FDR | Z | Cohen's d | p<sub>uncorr</sub> |
| Right DLPFC | 57 | 23 | 27 | 0.999 | 0.922 | 9 | 0.704 | 0.996 | 0.744 | 3.31 | 0.815 | 0.000 |

**Figure 2.**
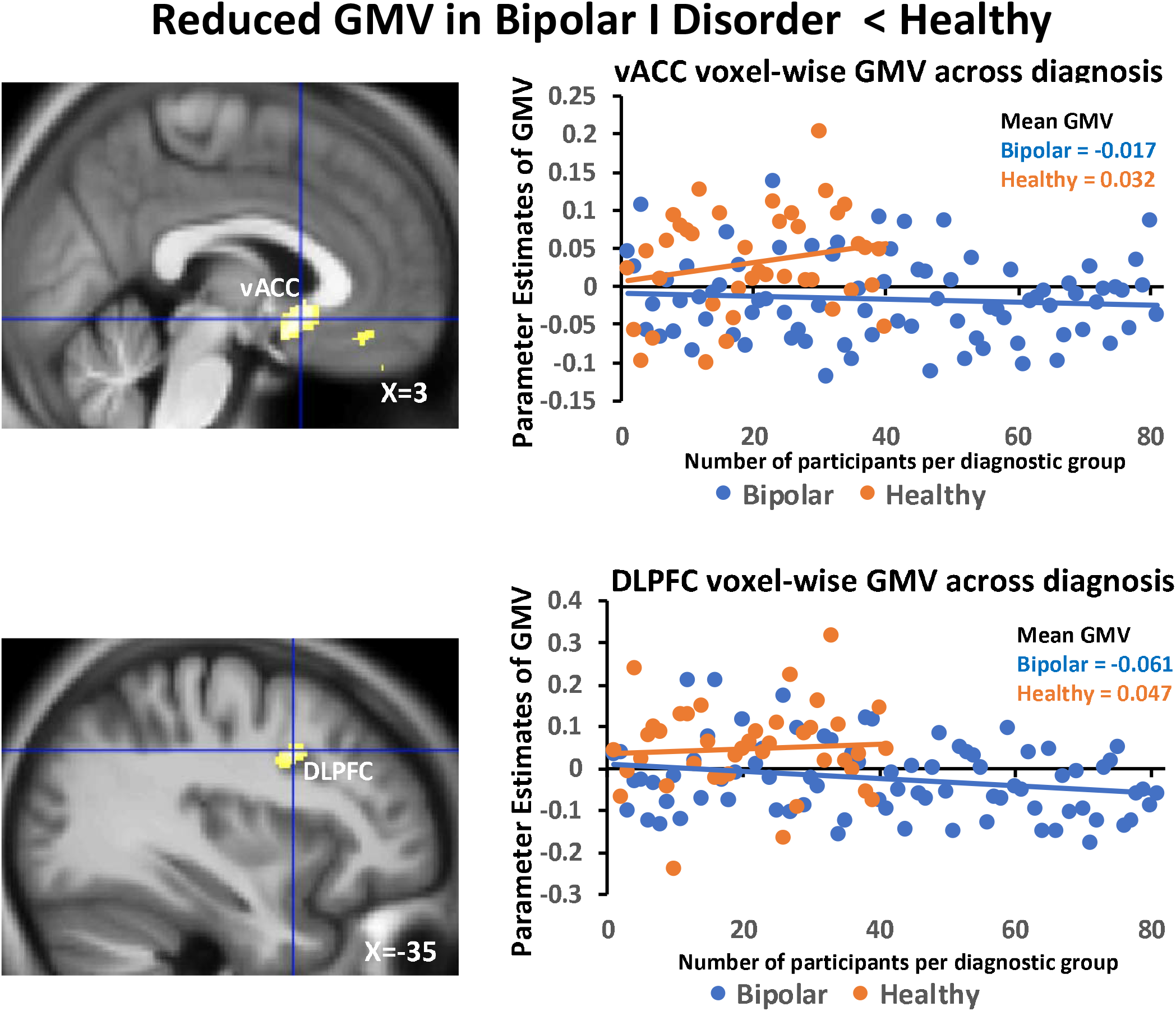
Frontolimbic volume-wise diagnostic group GMV differences. Whole-brain group difference of healthy vs bipolar GMV showing reduced volumes selectively localized in the vACC and ventral prefrontal cortex (top left image) and left DLPFC (top right image). The scatter plot shows the extracted individual GMV values shown to be relative more reduced in bipolar vs healthy in both vACC and DLPFC regions.

### Factorial ANOVA VBM Across Bipolar Suicide Attempt vs. Non-Attempt History

To further assess if measures of reduction in GMV within the anatomically defined frontolimbic ROIs might be more sub-regionally localized in relation to suicide attempt status, we conducted a full factorial ANOVA between the bipolar I individuals with a history of suicide attempts and bipolar I individuals without a history of suicide attempts. We first assessed suicide attempt history (i.e., a record of prior suicide attempt vs. no suicide attempt history) at the whole-frontolimbic/voxelwise level. Similar to the ROI analysis, we found that the main effect of suicide attempt history in terms of the magnitude of GMV reduction in vACC encompassing the rostral ACC/ventromedial prefrontal, and DLPFC were more prominent in those without suicide attempt history compared with those with a suicide attempt history at p < 0.001 uncorrected (**Figure 3A; Table 3B**). Note that the vACC was significantly reduced in bipolar versus controls, independent of suicide attempt history.

**Figure 3.**
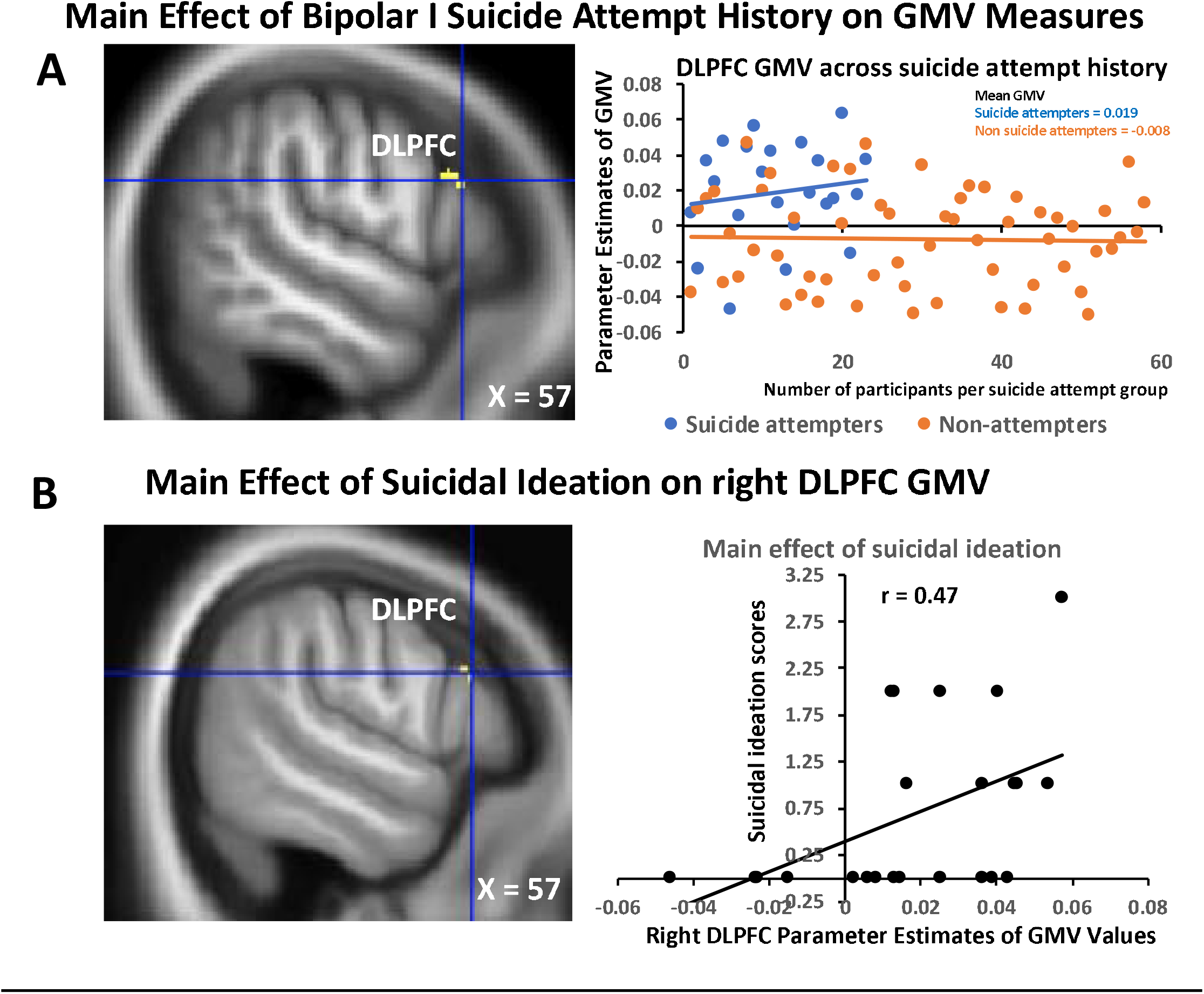
Voxel-wise GMV correlates for bipolar disorder suicide attempt history and suicidal state. A) shows GMV reductions in right DLPFC of bipolar participants without a history of suicide attempt relative to those with attempt history (i.e. main effect of suicide attempt) with the scatter plot showing the extracted individual GMV values illustrating the clustered group differences. B) the DLPFC region showing altered GMV in bipolar disorder in relation to suicide attempt history was found to correlate positively with/predict suicidal ideation in those with suicide attempt history.

We further assessed the relationship between GMV measures and current suicidal ideation scores and found that the observed individual variation in DLPFC GMV within the bipolar participants with suicide attempt history was positively correlated with HAMD-suicidal ideation scores (at r = .47) (**Figure 3B; Table 3C**). There was a negative correlation, albeit insignificant, between GMV measures in the DLPFC and suicidal ideation scores in bipolar individuals without a history of suicide attempt (at r = -.26) using the Pearson’s correlation in SPM12 at p < 0.001. Together, our findings that variability in reduction of GMV measures in the DLPFC selectively correlated with variability in suicidal ideation at the time of scanning in the bipolar cohorts with a suicide attempt history (r = .47), but inversely and more weakly in bipolar cohort with no history of suicide attempt (r = -.26). This finding suggests a possible non-linear relationship between observed regional GMV measures and suicidal behaviors.

## DISCUSSION

In this report, we tested the hypothesis that frontolimbic circuit anatomy would be altered in bipolar I disorder compared with healthy participants, and that these GMV differences would be associated with a history of a suicide attempt. We found that ACC and DLPFC regions that are known to be involved in integrating affective, inhibitory, and intero- and exteroceptive systems (Nauta 1971; Goldman-Rakic 1988; Critchley et al. 2004; Craig et al. 2009; Joyce & Barbas et al. 2018), which are relevant for high-level controls of affective processes including suicidal behavior, showed reduced GMV in bipolar I disorder vs. the healthy group. Further analyses showed a greater reduction in GMV in bipolar participants without a history of suicide attempts compared to those with a history of suicide attempt. This finding was contrary to our original hypothesis. It is however noteworthy that individual variability in suicidal ideation at the time of scanning only correlated with the observed DLPF reductions in GMV in the bipolar cohort with a suicide attempt history, suggesting the possibility that the GMV-related mechanisms associated with suicide attempt history are likely predictors of suicidal risk tendencies.

Here, frontolimbic GMV was found to be more reduced in the vACC ROI in bipolar disorder relative to healthy participants. This reduced vACC regional volume further exhibited a greater reduction in bipolar individuals without a history of suicide attempts compared to those with a history of suicide attempts. This localized vACC finding was in line with our DLPFC measures of GMV differences associated with suicide attempt history. The DLPFC results further showed that individual variations in our observed degree of frontolimbic DLPFC GMV reduction better predicted suicidal ideation in those with a history of suicide attempt than those without suicide attempt history and underscores the relevance of these anatomical regions in the inhibitory control of harmful thoughts and actions during a severe crisis (Schmaal et al. 2020). Taken together, our frontolimbic GMV bipolar diagnostic and suicide attempt history-related findings support a possible underlying frontolimbic mechanism that could contribute to the governing of adaptive and mal-adaptive excitatory/inhibitory balances in affective regulatory states (Joyce & Barbas 2018). Our results offer a potential window into the localized neuroanatomical repercussions for mood morbidity and suicide mortality. They suggest that the neuroanatomy for suicide attempt and ideation are likely more closely related than previously thought.

Several aspects of our findings deserve comment. *First*, suicide attempt history was not different between the bipolar depressed and manic cohorts. *Second*, the number of bipolar patients with suicide attempt histories was lower than those without suicide attempt history. As such, our multiple testing of a smaller sample with suicide attempt history can lead to medium-to-large effect sizes that could be inflated, especially in the whole-frontolimbic analysis that reflects highly localized GMV reductions related to suicidal behaviors.

*Third*, age of bipolar disorder onset was lower in the group without a history of a suicide attempt than in those that attempted suicide, whereas age at scanning was not significantly different between those with or without a suicide attempt history. As such, we cannot rule out the possibility that duration of bipolar illness (which was relatively longer in those without a suicide attempt history) may be more proximate to the degree of GMV reduction than our dichotomous measure of suicide attempt history. Given that the possible impact of longer illness duration is intertwined with the number of mood episodes across the bipolar illness lifespan, future studies need to better disentangle the interactive effects of the age of disease onset, mood state, and the number of depressed and manic episodes to better understand the impacts of these variables on GMV changes and associations with suicidal behavior. Additionally, while substance use/abuse status was an exclusion criterion, cannabis use was found to be higher in bipolar participants compared with healthy subjects, but not between manic vs. depressed bipolar individuals or between people with or without suicide attempter. Future studies need to better characterize possible effects of substance use as well as measures of impulsivity and socioeconomic stress, which were unaccounted for in the current study.

Fourth, the observed neurobiological phenotype in terms of vACC and DLPFC GMV associated with suicide attempt history reported here may be more proximal to the frequency of suicide attempts and the severity of suicide risk behavior rather than the dichotomous presence or absence of suicide attempt history. Furthermore, the observed findings may be, at least in part, driven by mood states because the participants in the depressed and manic cohorts were independent groups. This idea is consistent with our overall objective of using structural brain imaging measures — which are subservient to functional brain measures — to identify the underlying neuroanatomical mechanisms for suicide attempt risk across bipolar depressed and manic states, especially given the disproportionate number of suicides in bipolar disorder relative to other mental illnesses.

Our observation that frontolimbic GMV abnormalities in bipolar diagnosis and suicide attempt history can only be based on the presence of suicide attempt history since the frequency of suicide attempts were not explicitly analyzed continuously over time in the current study. Future studies using longitudinal approaches aimed at extending and replicating our findings by accounting for the number of prior suicide attempts across depressed and manic mood episodes documented over time in larger samples of the same individuals and age span will shed important light on the neuroanatomical changes driving suicidal behavioral risks.

In summary, our results underscore the anatomically relevant status of the targeted frontolimbic network’s structure (Hebb 1949; Nauta 1971; Goldman-Rakic 1988; Critchley et al. 2003; Damasio 2003; Jabbi et al. 2008; Craig 2009; Singer et al. 2009; Joyce & Barbas 2018) in the maintenance of affective states, including mood dysfunctions (Savitz & Drevets 2009; Goodkind et al. 2015; Wise et al. 2016) and their possible impacts on suicidal behavior. The observed frontolimbic GMV perturbations in terms of the most reduced volumes being present in bipolar participants, and that these reduced GMV locales were significantly correlated with measures of suicidal ideation in individuals with a suicide attempt history, is a striking demonstration of how neural systems serve to modulate the rupture of adaptive mood states which can increase suicidal behavioral risk tendencies. This line of research may inform the regional definition of molecular mechanistic biomarkers that can be used to advance novel mood disorder and suicide neurotherapeutics.

## Data Availability

This data is not currently available publicly but would be made available on request.

## Author Statement

Conception: MJ, WW, FN, DW, EL, MDB, CA, SS; Design and data collection: WW, MT, AG, CA; Data processing and statistical analysis: MJ, WW, JW; Manuscript Drafting: MJ; Contributions to writing of manuscript and other conceptual and intellectual contributions: WW, FN, DW, MT, AG, EL, MDB, CA, SS, CA, SS.

## Funding

This research is funded by R01MH080973

## Disclosures

M. Jabbi: None; W. Weber: None; J. Welge: None

F. Nery: Dr. Nery’s spouse is an employee of Eli Lilly & Co.

D. Fleck: None; A. Gable: None; M. Tallman: None

E. Lippard: receives research grant funding from Janssen pharmaceuticals.

M. DellBello: Dr. DelBello receives research support from NIH, PCORI, Acadia, Allergan, Janssen, Johnson and Johnson, Lundbeck, Otsuka, Pfizer, and Sunovion. She has also served as a consultant, on the advisory board, or has received honoraria for speaking for Alkermes, Allergan, Assurex, CMEology, Janssen, Johnson and Johnson, Lundbeck, Myriad, Neuronetics, Otsuka, Pfizer, Sunovion, and Supernus.

C. Adler: None

S. Strakowski: Dr. Strakowski chairs DSMBs for Sunovion as a Consultant, and has research grants with Janssen Pharmaceuticals and Alkermes.

---

**Supplementary Figure 1.**
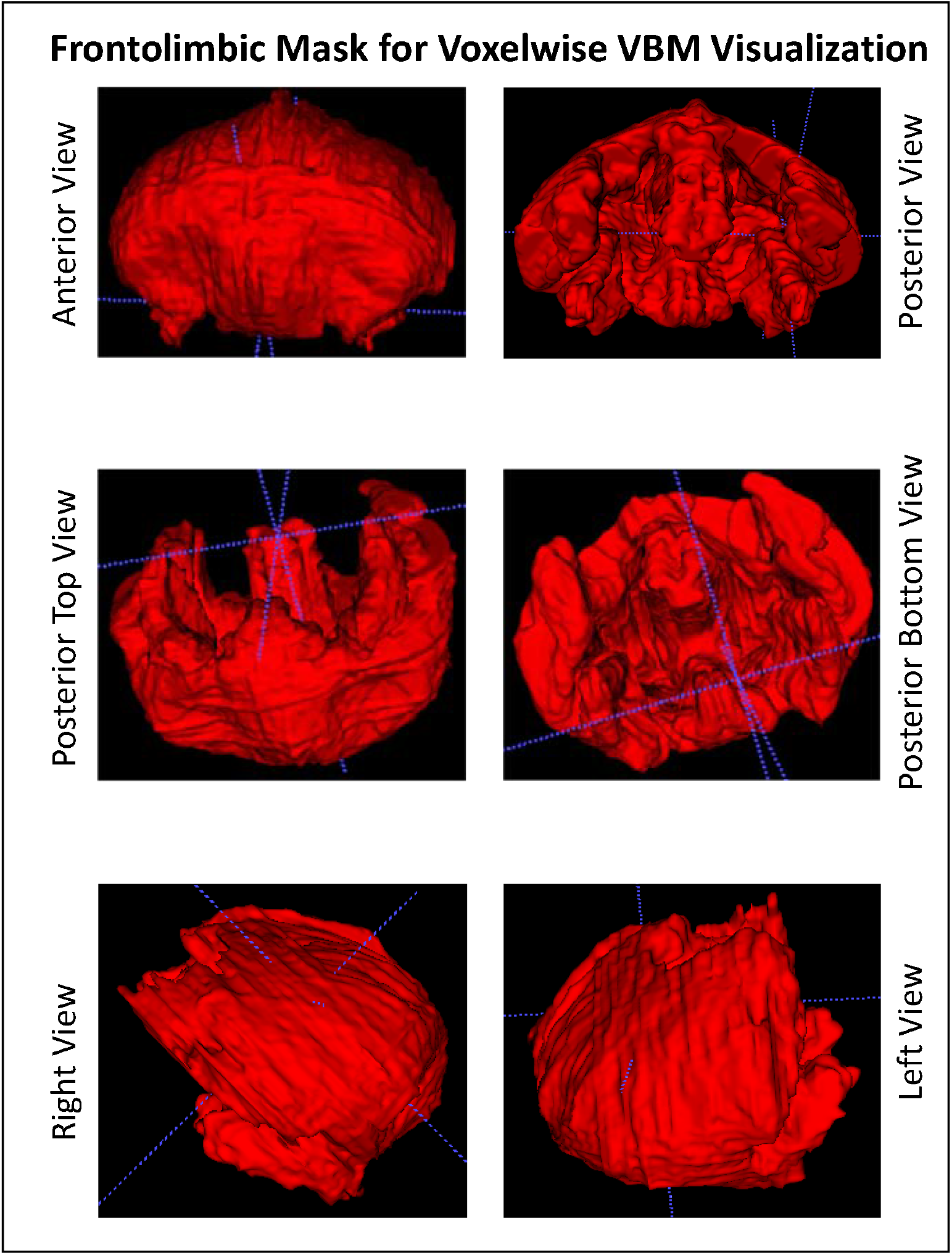
3-Dimensional Frontolimbic Mask used for visualization of all voxel-wise results of whole-brain VBM analysis. Note that the use of this mask does not affect the reported statistics in any way.

## Notes

### Competing Interest Statement

All authors have completed the ICMJE uniform disclosure form at www.icmje.org/coi_disclosure.pdf and declare: no support from any organisation for the submitted work; FN's spouse is an employee of Eli Lilly & Co. ECL receives research grant funding from Janssen pharmaceuticals. MDB receives research support from NIH, PCORI, Acadia, Allergan, Janssen, Johnson and Johnson, Lundbeck, Otsuka, Pfizer, and Sunovion. She has also served as a consultant, on the advisory board, or has received honoraria for speaking for Alkermes, Allergan, Assurex, CMEology, Janssen, Johnson and Johnson, Lundbeck, Myriad, Neuronetics, Otsuka, Pfizer, Sunovion, and Supernus. SS chairs DSMBs for Sunovion as a Consultant, and has research grants with Janssen Pharmaceuticals and Alkermes.

### Author Declarations

All participants received compensation for participating in the study, and the protocol was approved by the University of Cincinnati IRB.

### Summary of Updates

Most aspects of the previous submission have now been revised.

